# The socio-economic difference in the prevalence and treatment of diarrheal disease in children under five years across the geo-political zones in Nigeria

**DOI:** 10.1101/2021.10.30.21265706

**Authors:** Fatima Badmus Awoniyi, Subas Neupane

## Abstract

**Background:** Diarrhoeal disease is one of the leading causes of mortality among under-5 children globally and affects the low socio-economic population.

**Objectives:** This study aimed at evaluating the socioeconomic differences in the prevalence and its effect on the treatment of diarrheal disease in children in the different geopolitical zone in Nigeria.

**Methods:** Data of children under-5 from the 2018 Nigeria Demographic and Health Survey (NDHS) was used. A total of 30,068 women, that had at least a child under-5 years of age and answered diarrhoea specific questions were analysed for the prevalence, while data on 3885 children that reported symptoms of diarrhoea during the 2 weeks preceding the NDHS survey interview were analysed for treatment intervention offered during diarrhoea episodes. Logistic regression models adjusted for socio-demographic factors were used to study the association.

**Results:** The overall prevalence of diarrhoea was 12.9% with the highest prevalence in the North East (24.7%), among the poor (17.1%) and the children of uneducated mothers (16.4%). Compared to South-West region, children in North-East (AOR 4.64, 95% CI 3.90, 5.51), North-West (AOR 2.34, 95% CI 1.97, 2.78) and North Central (AOR 2.02, 95% CI 1.69, 2.42) had a high likelihood of having diarrhoea. Children from a poor household (AOR 1.49, 95% CI 1.31, 1.70) had more chance of having diarrhoea. Children in North-East (AOR 2.11, 95% CI 1.49, 3.01) and North-West (AOR 2.52, 95% CI 1.77, 3.60) were more likely to receive treatment in a health care facility and also had less likely to receive more amount of drink and food during diarrhoea compared to South-West region.

**Conclusions:** Diarrhoea prevalence is high in Nigeria with significant regional difference both in the prevalence and treatment of diarrheal disease which is also associated with household socio-economic status.

## BACKGROUND

Although childhood death due to diarrhoea among children under-5 has been reduced by 60% between 2000-2017 ^(1)^, it is one of the leading causes of mortality among under-5 children globally ^(2)^. The global incidence of the diarrheal disease still hits about 1.7 billion every year and it is still the second leading cause of death in children under-5 years old, causing an estimated 0.5 million under-5 deaths every year ^(3)^. According to WHO and Maternal and Child Epidemiology Estimation Group (MCEE 2018), it was estimated in 2017 that 424,000 under-5 deaths were due to diarrhoea globally, and 74,431 deaths in Nigeria ^(4)^. Despite being a global problem, locations with poor access to urgent health care, safe water and sanitation, and low-income or marginalised populations are disproportionately affected by a high burden of diarrhoea diseases ^(5)^. Globally, children from the poorest 20 per cent of households in their respective nations are at greatest risk of diarrhoea and are less likely to receive treatment than children from the richest quantile ^(1)^.

Diarrhoea is a risk factor for childhood undernutrition, wasting stunting and underweight and it has detrimental long term sequel associated with childhood growth impairment, causing growth faltering ^(6)^. Diarrhoea disease is associated with high mortality due to the pathophysiology of the disease, causing life-threatening dehydration, hence the need for adequate care during diarrhoeal episodes. Early diagnosis and appropriate treatment of diarrhoea are important in lowering the morbidity and mortality associated with diarrhoea ^(7)^.

Nigeria is the most populous country in Africa with a projected population of 392 million in 2050 ^(8)^. Despite the wealth of both human and natural resources endowed on Nigeria, globally, Nigeria remains one of the nations with widest inequality ^(9)^ with almost 100 million Nigerians living in abject or extreme poverty ^(9)^. Nigerian economic, political and educational resources are often shared across the six geopolitical zones, the major division in modern Nigeria. The six zones have not been entirely created solely based on geopolitical location, but rather states with similar cultures, ethnic groups, and common history were classified in the same zone. The creation of the zones was a result of the government’s need for the effective allocation of resources. ^(10)^ Several lines of evidence suggest that there is variability in the social-economic status of resident living in the different geopolitical zone of Nigeria ^(11)^

The household socioeconomic status influences the prevalence and treatment of diarrhoeal diseases ^(7)^. As the family is the first contact of a sick child, the welfare vis-a-vis the socio-economic status of the household is important in affecting the treatment interventions offered during childhood illness ^(7)^. However, to the best of our knowledge, none of the previous studies compared and analysed the prevalence and treatment of diarrhoea in the different geopolitical zone in relation to their socio-economic status. Thus, the level at which socio-economic status affect the treatment of diarrhoea disease in different regions of Nigeria needs to be evaluated to fill this literature gap. This study aimed at comparing the prevalence and the treatment of diarrheal disease in children in the different geopolitical zone in Nigeria based on their socioeconomic status and give recommendations towards improving the treatment across the different geopolitical zones in Nigeria. This will shed more light on the wealth distribution across the country and will be important in combating inequalities that occur within the country.

## MATERIALS AND METHOD

### Study Area, Study Design, And Data Source

The study area for this study was Nigeria. Nigeria comprises of 36 states which are grouped into 6 geopolitical zones namely: North-West NW, North-East NE, North Central NC, South-East SE, South-West SE and South-South SS. This study made use of the Nigeria Demographic and Health Survey (NDHS) 2018 data. The NDHS is nationally representative, population-based household surveys that monitor the change in demographics, reproductive health behaviours, attitudes and outcomes, child health, and socio-demographic characteristics of women and men of reproductive age ^(12)^. The DHS survey is a cross-sectional study and involved a stratified 2-staged cluster sampling procedure ^(13)^. Details of the NDHS sample design are explained elsewhere ^(13)^.

This study utilized data from the children recode of DHS dataset. Data were extracted for a total of 30,883 women who participated in the NDHS, 2018, that have at least a child under-5 year of age and answered diarrhoea specific questions. The respondents were women of reproductive age group (15-49) and are either permanent residents of the selected households or visitors who stayed in the households the night before the survey. The women were interviewed using the women’s DHS-7 women’s questionnaire.^(14)^ The women’s questionnaire included questions on background characteristics (age, education, religion, etc.), woman and child health, husband’s background and woman’s work, HIV/AIDS and other health issues ^(12)^. Information about diarrhoea symptoms in a child under-5 years of age in the household 2 weeks before the survey and information regarding the treatment interventions adopted, was based on the child’s health history information provided by the mother on the women’s questionnaire ^(12)^. Data on 3885 children that reported symptoms of diarrhoea during the 2 weeks preceding the NDHS survey interview were analysed for treatment intervention offered during diarrhoea episode.

### Measurement of variables

The variables extracted for analysis were based on evidence from previous studies ^(7; 10)^, available data and the research questions. The exposure variables for this study were factors that affect the prevalence of diarrhoea and the treatment interventions modalities adopted.

#### Diarrhoea related variables

The outcome variables are having diarrhoeal symptoms (Yes/No) and treatment interventions. Treatment interventions adopted for those who reported diarrhoeal symptoms including the use of Oral Rehydration Solution (ORS), zinc, antibiotic, health care intervention sought and nutrition. The “ORS Given” indicated the number of children with diarrhoea that received ORS. The variable was categorised and analysed as “Yes” or “No”. Nutrition during diarrhoea included the variables on use of zinc during diarrhoea episode, the amount offered to eat during diarrhoea episode and the amount offered to drink during diarrhoea episodes. “Given zinc” indicated the number of children with diarrhoea who received zinc. In the DHS data, the “don’t know” values were recategorized with the “No” category.

The variable on the amount of food and drink offered during diarrhoea episodes indicated the number of children in categories of the amount of drink and food given. The amount of drink offered during diarrhoea was categorised in the DHS data as “Nothing to drink”, “Much less”, “Somewhat less”, “About the same”, “More”, and “Don’t know”. In the descriptive analysis, we recategorized the variable as; “Less drink”, “About the same” and “More drink”. The variable on the amount of food offered during diarrhoea was measured in the DHS data as “Stopped food”, “Never gave food”, “Much less”, “Somewhat less”, “About the same”, “More”, and “Don’t know”. The variable was also recategorized as; “Less drink”, “About the same” and “More drink”. “Don’t know” values were recategorized with the “About the same”.

The variable on antibiotic use indicated the number of children with diarrhoea that received antibiotics. The ‘antibiotic used’ variable was created as a composite of “Given antibiotic pills or syrups” and the variable “Given antibiotic injection”. If either of the measurement was ‘yes’ then the new variable ‘antibiotic used’ was categorized as ‘yes’ and otherwise ‘no’.

The variable on the place of treatment sought indicated the place of first sought treatment. It was recategorized into four; public sector, private and other facility and no facility. The public sectors included government hospital, government health centre, government health post, public mobile clinic, public field worker and other public sectors. The private facility included a private doctor, private chemist, private mobile clinic, private field worker, and other private medical sectors. The other facility included: shops, traditional practitioner, market, itinerant drug seller, community-oriented resource person and others.

In the regression models, all the variables related to diarrhoeal treatment were dichotomized. The treatment facility was recategorized as “treatment facility sought” (consisted of the public sector, private and other facilities) and no “treatment facility”. Amount of the food and drink was recategorized as “given same or more food and drink” (consisted of “given more” and “given same” amount of food or drink) and given less food or drink.

#### Indicators of Socioeconomic Status

##### Wealth Index

The wealth index is the composite measure of a household’s cumulative living standard, and it is calculated based on the information concerning the household’s ownership of selected assets such as a television and car; dwelling characteristics such as flooring material; type of drinking water source; toilet facilities; and other characteristics that related to economic status ^(12)^. The wealth index in the DHS dataset was recoded as “poorest”, “poorer”, “Middle”, “Richer”, “Richest”. Based on evidence from previous research ^(15)^, the wealth index in this study was recategorized as “poor”, “middle” and “rich”. Other socioeconomic indicators used in the analysis are educational level of the mother, paternal educational level (no education, primary, secondary or higher), the literacy level of the mother, maternal occupational status, paternal occupational status, wealth Index, source of drinking water and sanitation facility.

### Statistical analysis

The data were analysed using SPSS version 26. The samples were weighted to adjust for the multistage stratified cluster sampling used in the NDHS.

For descriptive statistics, categorical variables were expressed as numbers and percentages. The prevalence of diarrhoea across the region and within the wealth-index quantiles and the maternal educational levels were calculated and presented as bar charts(Figure 1 and 2). The treatment options of diarrhoea within each region was assessed using Chi-squared test to study the regional difference. Binary logistic regression models were used to measures the association between the prevalence of diarrhoea and regions and the other independent variables on the indicators of socioeconomic status. The association between diarrhoea treatment of those who had diarrhoea symptoms and socioeconomic variables were studied using bivariate and multivariate regression models. The prevalence and treatment interventions for diarrhoea were studied across different geopolitical zones in Nigeria among groups classified based on their socioeconomic status using bivariate and multivariable logistic regression models. The odds ratio (ORs) and their 95% confidence intervals (CIs) was calculated from the regression models and adjusted for the socio-economic variables.

**Figure 1.**
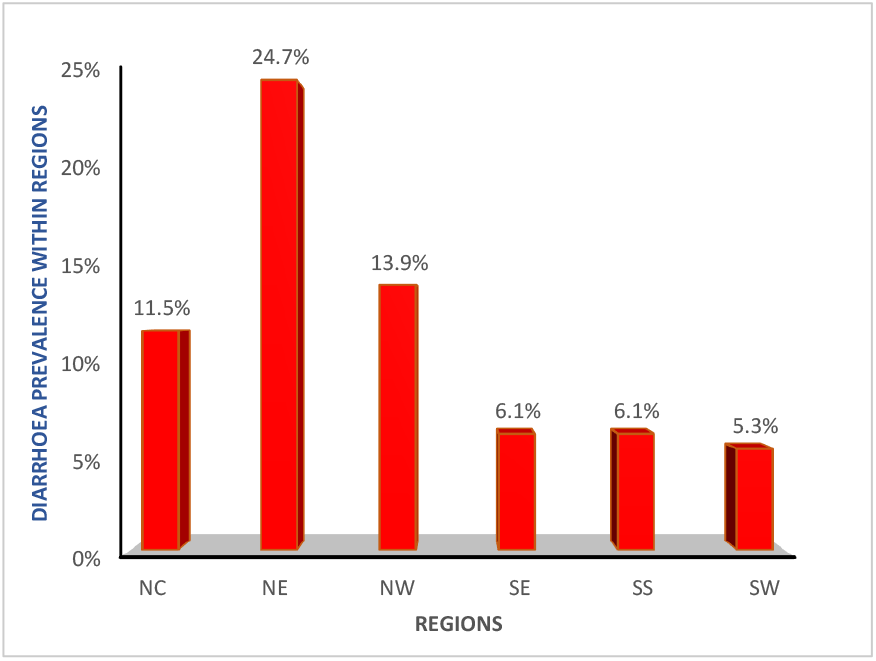
Prevalence of diarrhoea across the 6 geopolitical zones in Nigeria. Bivariate descriptive analysis (Crosstabulation) of Diarrhoea prevalence and Regions. The ‘Total’ represent only data of those that responded ‘yes’ to diarrhoea (3885) Diarrhoea prevalence is highest in NE with 24.7 % and lowest in SW with 5.3 % NC= North-Central, NE= North-East, NW= North-West, SE= South-East, SS= South-South, SW= South-West

**Figure 2a and 2b:**
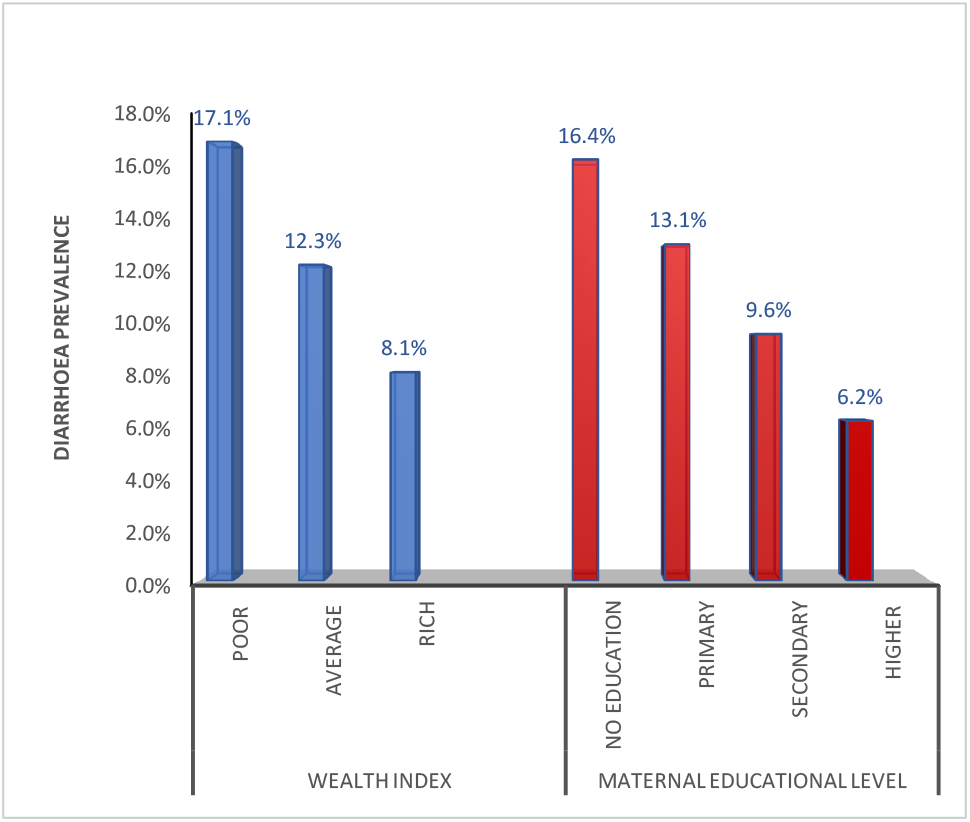
Prevalence of diarrhoea within the wealth index quantiles (2a) and Maternal educational levels (2b) Bivariate descriptive analysis (Crosstabulation) of Diarrhoea prevalence Socioeconomic Indicators(Wealth index and Maternal educational level) The ‘Total’ represent only data of those that responded ‘yes’ to diarrhoea (3884)

### Ethical consideration

Ethical considerations were made by DHS program during the survey. Procedures and questionnaires for standard DHS surveys were reviewed and approved by the Institutional Review Board (IRB). The DHS Program maintains strict standards for protecting the privacy of respondents and household members in all DHS surveys. Participants gave written/verbal consent to participate in the study ^(16)^.

## RESULT

### Descriptive Statistics

Table 1 presents the frequency distribution of the socio-demographic and socio-economic characteristics of the households. 35% of the respondents were resident of the North-West region while 8.9 % were from the South-South. The mothers’ age ranged from 15 to 49 years with about 70 per cent of the mothers within the ages of 30-34. 40% of the household had 2 children and about 33% had 3 or more children. Over 40% of the study population were poor. About 45% of the women had no education, 8.5% had higher education and 58.6% were not able to read at all. 37% of the respondents’ partners had no education and just 15% of the partners had higher education. Over half (67%) of the women surveyed reported to be working and the majority of their partners (96%) were also working. Just over half of the household had access to an improved source of drinking water (51%) and improved sanitation (51.3%).

**Table 1.** Univariate descriptive analysis of socio-demographic and socio-economic variables.

|  |  |  |  |
| --- | --- | --- | --- |
|  |  | Frequency N=30068 | Percent |
| Age of mother | <20 years | 1255 | 4.2 |
|  | 30-34 | 21241 | 70.6 |
|  | 35 years or more | 7572 | 25.2 |
| No Children under 5 in Household | 1 child | 7936 | 26.4 |
|  | 2 children | 12198 | 40.6 |
|  | 3 or more children | 9934 | 33 |
| Highest educational level of the mother | No education | 13669 | 45.5 |
|  | Primary Education | 4499 | 15 |
|  | Secondary education | 9345 | 31.1 |
|  | Higher education | 2556 | 8.5 |
| Husband's/ partner's educational level | No education | 10750 | 37.3 |
|  | Primary Education | 3956 | 13.7 |
|  | Secondary education | 9775 | 33.9 |
|  | Higher education | 4345 | 15.1 |
| Literacy level of the mother | Cannot read at all | 17623 | 58.6 |
|  | Able to read | 12445 | 41.4 |
| Maternal occupational status | Not working | 9742 | 32.4 |
|  | Working | 20327 | 67.6 |
| Husband's/ partner's occupational status | Not working | 1004 | 3.5 |
|  | Working | 27823 | 96.5 |
| Source of drinking water | Improved sanitation | 15373 | 51.1 |
|  | Unimproved facility | 11141 | 37.3 |
|  | No facility | 3385 | 11.3 |
| Sanitation facility | Improved sanitation | 15330 | 51.3 |
|  | Unimproved sanitation | 7747 | 25.9 |
|  | No facility | 6821 | 22.8 |
| Wealth index | Poor | 13176 | 43.8 |
|  | Average | 6173 | 20.5 |
|  | Rich | 10719 | 35.7 |
| Region | North Central | 4113 | 13.7 |
|  | North East | 5479 | 18.2 |
|  | North West | 10756 | 35.8 |
|  | South East | 3050 | 10.1 |
|  | South | 2689 | 8.9 |
|  | South West | 3983 | 13.2 |
| Diarrhoea Prevalence | No diarrhoea | 26184 | 87.1 |
|  | Yes | 3884 | 12.9 |
| N= Total number of respondents that answer questions on diarrhoea |  |  |  |
| *There are some missing values. So, the total number for N for the variables may not be up to (30068)100% |  |  |  |

### Diarrhoea Prevalence

About 13% (3884) reported having a child with diarrhoea within 2 weeks before the survey. The prevalence of diarrhoea was highest in the North-East of Nigeria with a prevalence of 24.7%. The southern regions had a significantly lower prevalence of diarrhoea with the South-West region having a prevalence of 5.3% (Figure 1). The prevalence of diarrhoea was also highest among the poor (17.1%) and the children of uneducated mothers (16.4%) but lowest among the rich (8.1%) and among children of mothers with higher educational level (6.2%).(Figure 2a and 2b)

### Treatment of Diarrhoea

Data on those children that had diarrhoea (n=3885) were further analysed based on the diarrhoea treatment interventions adopted.(Table 2) In North-West, 28.8% of the children had treatment in a public facility, while only 15.2% had so in South-East. A higher percentage of the children in the South-East and South-South received health care in private health facilities with 41.8% and 47.9% respectively, while only 29.2% in South-West sought care in a private facility. A large proportion of household in the North-Central region (42.6%) and South-West region (40%) did not attend health facility. 50.9% of children were offered ORS in the South-West while more than 60% of the children in North-Central and North-East had no ORS. Although, there is an obvious less use of antibiotics across all the regions, a significantly high proportion of children in South-West Nigeria (89.2%) were not offered antibiotics during the course of the diarrhoea. However, 39.2% of children in the North-Central had antibiotic treatment. 67% of children in North-East received less than their regular amount of food and over 60% of children in North-West were offered less than their regular amount of food and drink. The same amount of food and drink were offered to over 55% of children in the South-West. Use of zinc was low across all the regions. However, the North West region had a relatively higher proportion (42%) of children that received zinc, while 78.2% of children from the North East had no zinc offered.

**Table 2.**
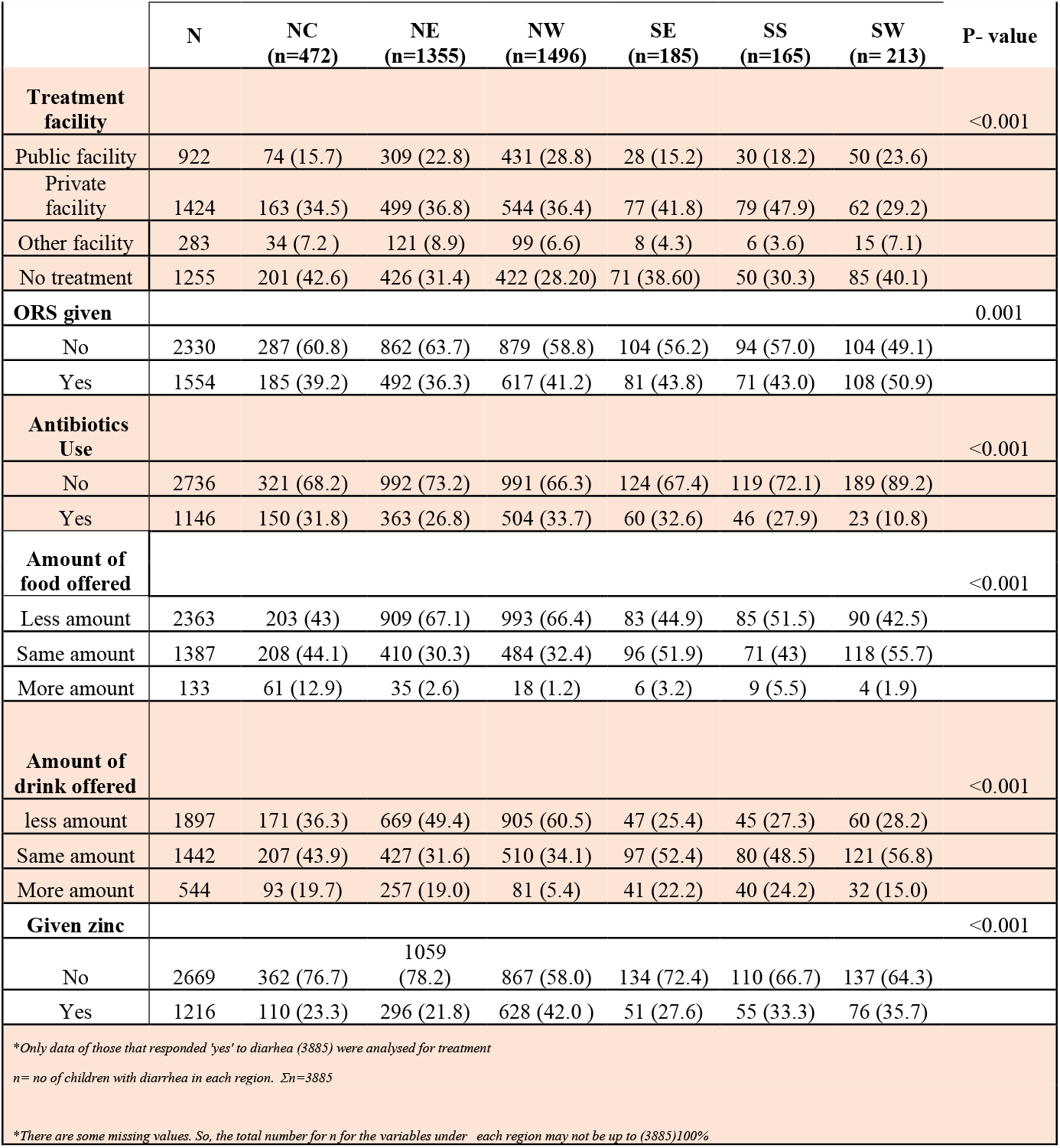
Treatment interventions for diarrheal across different Regions Regions(n, % within region)

### Association of the prevalence of diarrhoea with regions socio-economic status

Table 3 present the association of the diarrhoea prevalence with the region and the socioeconomic status of the household. Odds ratio were adjusted for all socio-demographic factors used. The adjusted model shows that the North East region had 4.6 times higher likelihood of having diarrhoea (AOR 4.64, 95% CI 3.90, 5.51) compared to South-West region where the prevalence rate was the lowest. Similarly, other Norther regions (North-West and North-Central) also had a higher likelihood of having diarrhoea compared to the South-West region (AOR for NW region =2.337, 95% CI 1.97, 2.78) and (AOR for NC region = 2.02, 95% CI 1.70, 2.42).

**Table 3.** Binary logistic regression of prevalence of diarrhoea with the regions and the socio-economic indicators.

|  |  |  |  |  |  |  |  |
| --- | --- | --- | --- | --- | --- | --- | --- |
|  |  | Crude Odds Ratio |  |  | Adjusted Odds Ratio |  |  |
|  |  | OR | 95% C.I. |  | OR | 95% C.I. |  |
|  |  |  | Lower | Upper |  | Upper | Lower |
| Regions | NC | 2.30 | 1.94 | 2.72 | 2.02 | 1.69 | 2.42 |
|  | NE | 5.82 | 5.00 | 6.78 | 4.64 | 3.90 | 5.51 |
|  | NW | 2.87 | 2.45 | 3.32 | 2.34 | 1.97 | 2.78 |
|  | SE | 1.14 | 0.93 | 1.40 | 1.11 | 0.90 | 1.38 |
|  | SS | 1.16 | 0.94 | 1.43 | 1.07 | 0.86 | 1.34 |
|  | SW (Ref) | 1.00 |  |  | 1.00 |  |  |
| Wealth Index | Poor | 2.34 | 2.15 | 2.54 | 1.49 | 1.31 | 1.70 |
|  | Average | 1.59 | 1.44 | 1.77 | 1.20 | 1.07 | 1.36 |
|  | Rich(Reference) | 1.00 |  |  | 1.00 |  |  |
| Maternal Educational level | No education | 2.97 | 2.51 | 3.51 | 1.58 | 1.24 | 2.01 |
|  | Primary | 2.28 | 1.90 | 2.74 | 1.71 | 1.37 | 2.14 |
|  | Secondary | 1.62 | 1.36 | 1.93 | 1.57 | 1.29 | 1.90 |
|  | Higher(Reference) | 1.00 |  |  | 1.00 |  |  |
| Paternal Educational level | No education | 1.54 | 1.39 | 1.72 | 0.80 | 0.70 | 0.93 |
|  | Primary | 1.09 | 0.95 | 1.24 | 0.77 | 0.66 | 0.90 |
|  | Secondary | 0.94 | 0.84 | 1.05 | 0.84 | 0.74 | 0.95 |
|  | Higher(Reference) | 1.00 |  |  | 1.00 |  |  |
| Maternal Literacy level | Not able to read | 1.74 | 1.61 | 1.87 | 0.95 | 0.83 | 1.08 |
|  | Able to read part or whole sentence (Reference) | 1.00 |  |  | 1.00 |  |  |
| Maternal occupation | Not working | 1.06 | 0.99 | 1.14 | 0.89 | 0.82 | 0.96 |
|  | Working (Reference) |  |  |  |  |  |  |
| Paternal occupation | Not working | 0.59 | 0.47 | 0.74 | 0.53 | 0.42 | 0.67 |
|  | Working (Reference) | 1.00 |  |  | 1.00 |  |  |
| Source of drinking water | Improved facility (Reference) | 1.00 |  |  | 1.00 |  |  |
|  | unimproved facility | 1.51 | 1.41 | 1.62 | 1.03 | 0.95 | 1.12 |
|  | no facility | 1.00 | 0.89 | 1.13 | 0.89 | 0.78 | 1.01 |
| Sanitary Facility | Improved facility(Reference) | 1.00 |  |  | 1.00 |  |  |
|  | unimproved facility | 1.48 | 1.37 | 1.60 | 1.04 | 0.94 | 1.14 |
|  | no facility | 1.12 | 1.03 | 1.23 | 0.93 | 0.83 | 1.04 |

Among socioeconomic indicators, the children from the poor household had a 1.5-fold higher likelihood of having diarrhoea (AOR 1.49, 95% CI 1.31, 1.70) compared to rich households. Furthermore, the odds of having diarrhoea were about 2 times more among children whose mother had no education (AOR 1.6, 95% CI:1.24, 2.01), primary education (AOR 1.71, 95% CI 1.37, 2.14) and secondary education (AOR 1.57, 95% CI 1.29, 1.90) compared to higher education. Children whose mothers were not working had about 20% less chance of having diarrhoea (AOR 0.89 95% CI: 0.623, 0.96). The prevalence of diarrhoea was lower by about 50% in children whose fathers were working (AOR 0.59, 96% CI 0.47, 0.736) compared to not working partner.

### Regional difference in diarrhoeal treatment

Table 4 illustrate the regional difference in diarrhoeal treatment. Compared to South-West region, the adjusted odd ratio in all other regions shows a higher likelihood of receiving antibiotics among children who had diarrhoea with the highest odds in the North-Central region (AOR 4.65, 95% CI 2.82, 7.64). Similarly, compared to the South-West region, the children in other regions had a lesser likelihood of ORS treatment for diarrhoea (Model 1). Treatment facility sought was more likely for children in North-East (AOR= 2.11, 95% CI 1.4 9, 3.01), North-West (AOR= 2.52, 95% CI 1.77, 3.60) and South-South (AOR= 1.61, 95% CI 1.02, 2.57) regions compared to South-West region. However, children in North-West had twice the likelihood of receiving zinc (AOR 2.12, 95% CI 1.49, 3.01). Giving children more amount of drink and food was less likely in North-East (drink, AOR 0.37, 95% CI 0.26, 0.53 and for food, AOR 0.38, 95% CI 0.27, 0.53) and North-West (drink, AOR 0.23, 95% CI 0.16, 0.33 and for food (AOR 0.42, 95% CI 0.30, 0.59) compared to South-West region.

**Table 4.** Multivariable analysis using binary logistic regression of the regions and diarrhoea treatment

|  | NC |  | NE |  | NW |  | SE |  | SS |  |
| --- | --- | --- | --- | --- | --- | --- | --- | --- | --- | --- |
|  | COR | AOR | COR | AOR | COR | AOR | COR | AOR | COR | AOR |
|  | OR ( 95 % CI) | OR ( 95 % CI) | OR ( 95 % CI) | OR ( 95 % CI) | OR ( 95 % CI) | OR ( 95 % CI) | OR ( 95 % CI) | OR ( 95 % CI) | OR ( 95 % CI) | OR ( 95 % CI) |
| Give ORS | 0.62 (0.45,0.86) | 0.80 (0.56,1.15) | 0.55 (0.41,0.74) | 0.75 (0.54,1.05) | 0.68 (0.51,0.90) | 0.99 (0.70,1.38) | 0.75 (0.50,1.11) | 0.81 (0.53,1.24) | 0.73 (0.49,1.10) | 0.87 (0.56,1.36) |
| Antibiotics used | 3.82 (2.38,6.13) | 4.65 (2.82,7.64) | 2.98 (1.91,4.67) | 3.51 (2.16,5.70) | 4.15 (2.66,6.47) | 4.85 (2.99,7.86) | 3.97 (2.34,6.74) | 4.53 (2.61,7.88) | 3.14 (1.81,5.44) | 4.05 (2.28,7.20) |
| Treatment facility sought | 0.91 (0.65,1.26) | 1.22 (0.85,1.75) | 1.46 (1.09,1.97) | 2.11 (1.49,3.01) | 1.71 (1.27,2.30) | 2.52 (1.77,3.60) | 1.07 (0.72,1.60) | 1.17 (0.76,1.80) | 1.54 (1.00,2.37) | 1.61 (1.02,2.57) |
| Zinc used | 0.55 (0.38,0.78) | 0.79 (0.54,1.15) | 0.50 (0.37,0.68) | 0.80 (0.56,1.14) | 1.30 (0.97,1.76) | 2.12 (1.49,3.02) | 0.68 (0.45,1.05) | 0.81 (0.51,1.27) | 0.91 (0.59,1.39) | 1.07 (0.68,1.70) |
| Given same or more drink | 0.70 (0.49,0.99) | 0.71 (0.49,1.03) | 0.41 (0.30,0.56) | 0.37 (0.26,0.53) | 0.26 (0.19,0.36) | 0.23 (0.16,0.33) | 1.16 (0.74,1.81) | 1.26 (0.79,2.02) | 1.04 (0.66-,1.64) | 1.08 (0.67,1.75) |
| Given same or more food | 0.98 (0.70,1.35) | 1.01 (0.71,1.43) | 0.36 (0.27,0.49) | 0.38 (0.27,0.53) | 0.37 (0.28,0.50) | 0.42 (0.30,0.59) | 0.90 (0.61,1.34) | 1.04 (0.68,1.57) | 0.69 (0.46,1.03) | 0.67 (0.43,1.04) |

COR= Crude Odds Ratio, AOR= Adjusted Odds Ratio, OR= Odds Ratio CI= Confidence Interval , NC= North-Central, NE= North-East, NW = North-West, SE= South-East, SS= South-South
Reference = South-East (OR= 1)

## DISCUSSION

This study investigated the prevalence and the treatment of diarrhoea across the different geopolitical zones in Nigeria based on their socioeconomic status. Despite evidence demonstrating the importance of the use of ORS in the management of diarrheal ^(17)^ and of the initiation of Global Action For the Prevention and Treatment of Pneumonia and Diarrhoea, GAPPD which aims toward reducing diarrhoea mortality and improving the treatment for diarrhoea, there is still a high prevalence of diarrhoea in Nigeria with a significant regional discrepancy in the prevalence and optimal treatment. It was observed that compared to the rich households, children from poor households had a higher likelihood of having diarrhoea and children from the Northern region (North-East and North-West) were more likely to receive treatment in a health care facility and also less likely to receive more amount of drink and food during diarrhoea compared to South-West region.^(18)^

In this study, the overall prevalence of diarrhoea was found to be 12.9%. A high prevalence of diarrhoea was also observed in Southern Ethiopia 13.6 % ^(19)^, Northwest Ethiopia 14.5% ^(20)^ and in Ghana 15.4% ^(21)^. However, this finding was lower than the studies in India 25.2 %^(22)^, Ethiopia 22% ^(18)^, Burundi 32,6%^(23)^, Somalia and Eastern Ethiopia 27.3% and in Yemen 29.09%^(24)^. The high prevalence of diarrhoea in these countries including Nigeria may be attributed to their socio-economic status. Diarrheal illness is more predominant in low - and middle-income countries due to low socio-economic status^(25)^. Although 62% of the global under-5 population resides in the low and lower-middle-income countries, they account for more than 90% of global diarrhoea death^(26)^.

On the regional level, this study found Northern regions to have a significantly higher prevalence (24.7% in the North-East) compared to the southern regions (5.3% in South-West). The observed regional differences are in line with an earlier study ^(27)^ which showed a regional discrepancy in the distribution of diarrhoea with an increasing prevalence of the diarrhoeal disease among infants in the Northern part of Nigeria than in the South with a prevalence of 21.1% in the North-West^(27)^. A similarly high prevalence was observed by Omole et.al, with a prevalence of 30.29% in North-West Nigeria^(28)^. This regional difference may be a reflection of the underlying inequality that exists across the geopolitical zones in Nigeria. Nigeria remains one of the nations with the widest inequality with almost 100 million Nigerians living in abject or extreme poverty ^(9)^.

The result from this study also demonstrated a significant association between wealth index and diarrhoea prevalence with children from the poor household having twice the chance of having diarrhoea compared those from rich households. This is in line with studies in North-East Ethiopia,^(29)^ and Ghana.^(30)^and maybe due to poor access to environmental facilities, poor sanitary conditions and poor access to safe drinking water in the households. The 2016 World Bank report has shown a wide poverty disparity between the Northern and the South region of Nigeria ^(31)^. Thus, it is reasonable to say that the regional difference in the prevalence of diarrheal may be due to the low socio-economic status of the households in the northern regions.

Furthermore, this study found that the regional disparity extends to the treatment interventions during diarrhoea episodes. It is quite surprising that, despite the higher chance of receiving health care from the health facility, children in the Northern region (North-West and North-East) were less likely to receive an adequate amount of food and drink during diarrhoea episodes compared to the South-West. The higher likelihood to seek health care treatment in the Northern region contrasts the findings in South-West Ethiopia ^(32)^ and the 2016 World Bank report on Nigerian poverty work program which suggested disproportionately lesser access of the Northern region to basic infrastructures such as health facilities compared to the South zones^(31)^. A likely explanation for increase seeking of health at a health facility in the Northeast and North-West despite the level of poverty may be due to the presence of severe and life-threatening forms of diarrhoea which necessitate health facility treatment. Evidence of increased malnutrition in the Northwest and Northeast has been illustrated in pieces of literature^(10; 33)^. Malnutrition may predispose children to more severe forms of diarrhoea.

The observed suboptimal feeding received by children of the North-East and North-West in this study during diarrhoea episodes is in contrast to the study in Pakistan ^(34)^ and Peru^(35)^. With the previous evidence of malnutrition in the Northern states, this suboptimal feeding may be a continuum of the poor feeding habits that predisposed them to malnutrition. Malnutrition is a complication and also a risk factor for diarrhoea disease. Sufficient nutrition through continued feeding during diarrhoea episodes reduces the severity, enhance recovery and improve resistance to future diarrheal episodes. ^(36)^

Although the use of ORS was not optimal across the whole country, the children in the Northern region had relatively lower use of ORS compared to the South-West. This is in line with the findings in the study in Cameroon^(37)^ and by Reiner et al.,^(38)^ that reported geographical inequalities in ORS administration in low-and-middle-income countries such as Nigeria. The low use of ORS may be due to the poor economic status of the household which can be supported by findings in Eastern Ethiopia^(39)^. Other evidence showed the suboptimal use of ORT by the parents of children living in poverty^(40)^. It appears the major regional difference in the treatment and prevalence of diarrhoea in Nigeria can be arguably attributed to household poverty.

DHS surveys are cross-sectional surveys and routinely gathers 2-week point prevalence rates. Since the information of the child’s illness was from the mother, there is a chance of respondent’s recall and reporting bias on the interventions offered during a diarrheal episode that occurred within the 2 weeks preceding the survey. This could have possibly influenced the results. However, this the methodology in this study was an in-depth analysis of regional difference that gives a relative glimpse of regional discrepancy in Nigeria.

## Conclusions

There is a regional difference in the prevalence and treatment of diarrheal disease in children under-5 in Nigeria and this is affected by household poverty. The government should make policies that will improve household poverty especially in the Northern part of Nigeria and also formulate policies towards ensuring equity in the distribution of wealth and health care services related to diarrhoea management across the country.

## Data Availability

All data produced in the present study are available upon reasonable request to the authors

https://dhsprogram.com/

## Acknowledgement

The authors are grateful to DHS, ICF International, Rockville, Marylands, USA for providing all the 2018 NDHS data for this analysis.

